# Cell-Free DNA GWAS Reveals Importance of p.Arg206Cys in DNASE1L3 for Non-Invasive Testing

**DOI:** 10.1101/2024.08.15.24312005

**Authors:** Jasper Linthorst, Michel Nivard, Erik A. Sistermans

## Abstract

Properties of cell-free DNA (cfDNA) are intensely studied for their potential as non-invasive biomarkers. We explored the effect of common genetic variants on concentration and fragmentation properties of cfDNA using a GWAS based on low-coverage whole genome sequencing data of 140.000 Dutch Non-Invasive Prenatal Tests (NIPT). Our GWAS detects many genome-wide significant loci, functional enrichments for Phagocytes, Liver, Adipose tissue, Macrophages and genetic correlations with autoimmune and cardiovascular disease. A common (7%) missense variant in *DNASE1L3* (p.Arg206Cys), strongly affects all cfDNA properties. It increases the size of fragments, lowers cfDNA concentrations, affects the distribution of cleave-site motifs and increases the fraction of circulating fetal DNA during pregnancy. For the application of NIPT, and potentially other cfDNA-based tests, this variant has direct clinical consequences as it increases the odds of inconclusive results and impairs the sensitivity of NIPT by causing predictors to overestimate the fetal fraction.

**HIGHLIGHTS:**

- Common variants affect properties of plasma cell-free DNA
- p.Arg206Cys in *DNASE1L3* strongly affects the size of cell-free DNA fragments
- Fragmentomics-based fetal fraction predictors are affected by p.Arg206Cys
- Genetics behind cfDNA overlaps with autoimmune and cardiovascular diseases

## INTRODUCTION

Plasma cell-free DNA (cfDNA) are extracellular DNA fragments present in blood. These fragments predominantly originate from the degradation of dead cells, which results in non-randomly fragmented cfDNA fragments that circulate in plasma. Most cfDNA originates from apoptotic hematopoietic cells, but a substantial amount originates from other tissues. Changes in the pool of cfDNA can be observed in pathological as well as physiological conditions such as growth of a tumor or placenta, tissue damage, presence of a graft and graft rejection, microbial infection, or the innate immune response against an infection. Plasma cfDNA can easily be obtained from blood, without the risks that are involved with invasive procedures. This has enabled the quick adoption of clinical applications such as non-invasive prenatal testing (NIPT) and ‘liquid biopsies’ in oncology and transplantation monitoring.

Across these disciplines, applications face very similar challenges, such as distinguishing fragments of interest from a background of highly similar cfDNA and making inferences about the cell-type composition of the cfDNA pool. To do so, researchers have explored and made use of the fragmentation properties of cfDNA molecules, a study referred to as ‘fragmentomics’^1^. These properties include, but are not limited to: cfDNA fragment sizes, 5’ end-patterns, ‘jaggedness’^2^, ‘preferred ending-sites’^3^, ‘window-protection scores’^4^ and many others. The predictive potential of these features is attributed to the fact that the non-random fragmentation patterns of cfDNA fragments reflect the epigenetic and transcriptomic footprints of the cells from which they originate^1^.

To better understand the mechanisms behind fragmentomics, a model of cfDNA biology has been conceived, which mainly focuses on the nucleases involved in the degradation of apoptotic cells, and revolves around three main nucleases, *DFFB*, *DNASE1*, and *DNASE1L3*^5^. Briefly, during apoptosis, *DNASE1L3* and *DFFB* (also known as DNA fragmentation factor B or caspase-activated DNase) first intracellularly cleave chromatin in the internucleosomal linker region into large oligo-nucleosomal DNA fragments. After the initial intracellular cleavage, the apoptotic cell breaks apart into apoptotic bodies, which are eventually cleared by phagocytosis of macrophages. This process predominantly takes place in the liver and spleen. The nucleosome-bound DNA fragments that during this process are released into the bloodstream are further degraded by secreted circulating enzymes. DNASE1L3 and DNASE1 account for most of the direct DNase activity in blood, but other circulating enzymes have been shown to contribute to this process by releasing nucleosomes from apoptotic bodies, such as the Factor VII activating protease (HABP2 or FSAP)^6,7^ and Factor H^8^. Neither *DNASE1*, *DNASE1L3* or *DFFB* has a simple consensus cleavage sequence, but *DFFB* has been shown to prefer cleavage of DNA at the center of a 8bp palindromic Purine(R)/Pyrimidine(Y) sequences (5’-RRRY|YRRR-3’)^9^.

Besides diagnostic purposes, plasma cfDNA in vivo is not merely a byproduct of cell-death. It has a pro-inflammatory effect, which plays an important role in the innate immune response^10,11^. As part of the innate immune response, Neutrophils, the most abundant leukocyte in the bloodstream^12^, but also Eosinophils and Basophils, can produce Extracellular DNA traps, which are often referred to as Neutrophil Extracellular Traps (NETs). NETs are long stretches of chromatin and mitochondrial-derived DNA, coated with anti-microbial proteins, which are intended to trap and kill pathogenic microbes^13^. NETs are mostly the result of a non-apoptotic cell-death mechanism, referred to as NETosis. It is currently unclear how much cfDNA is attributable to NETs^14^, specifically under physiological conditions^15^, but total plasma cfDNA concentration measures have been used and validated as surrogate biomarkers for the detection of NETs and NETosis in disease^16^. NETs provide a functional link between cfDNA, inflammation, and coagulation^17–19^, which is often referred to as immunothrombosis, and is believed to play a role in many other diseases and pathological conditions, such as cardiovascular disease, sepsis, atherosclerosis, thrombosis and cancer.

There is also considerable evidence that cfDNA and NETs play an important role in the development of autoimmunity^14,11,20^. Additionally, impaired clearance of both inter- and intracellular DNA has been shown to cause or increase the risk of different autoimmune diseases. This mechanism is supported by mouse models and patients with loss-of-function variants in nuclease genes. Rare variants in *TREX1*^21,22^, *SAMHD1*, *ADAR1*, *DNASE2*^23^, *DNASE1*^24^ and *DNASE1L3*^25–27^ are all causally associated with different autoimmune diseases. A common missense variant in *DNASE1L3*, here referred to as R206C, was recently identified as the causal variant responsible for the signal on 3p14.3 (often wrongfully assigned to the neighboring *PXK* gene) in Systemic Lupus Erythematosus (SLE)^28,29^, and also underlies GWAS associations from Rheumatoid Arthritis (RA)^30^ and Systemic Sclerosis (SS)^31^. Additionally, recent work has shown that antibody-mediated inhibition of *DNASE1L3* function is a non-genetic mechanism which contributes to SLE pathogenicity^32^.

Most of our current knowledge stems from studies in mice, where nuclease genes were knocked-out in order to explain the characteristic patterns of cfDNA. So far, this has resulted in a model for cfDNA generation which translated well to humans and has provided crucial insights into many aspects of fragmentomics^5^. Despite this model, there are still many open questions concerning cfDNA, which are key to unlocking new insights into cfDNA diagnostics, autoimmunity and immunothrombosis^15^.

In this paper, we aim to contribute to these open questions by performing several genome-wide association studies (GWAS) on different fragmentation and concentration properties of cfDNA. For this, cfDNA sequencing data is used from approximately 140.000 pregnant women enrolled in the TRIDENT-2 study which implemented NIPT in The Netherlands^33^. Data from NIPT analyses performed in Amsterdam were used to impute and investigate the association between common variants and cfDNA traits, calculated and measured from the same data.

Although a previous study using NIPT collected in China already showed the potential for this source of data to perform GWAS^34^, it wasn’t used to study the biological properties of cfDNA. Our study using NIPT data collected in The Netherlands is furthermore characterized by a higher per sample sequencing depth and the ability to study the size of cfDNA fragments due to the use of paired-end sequencing technology.

We derive estimates for the common SNP heritability of these traits, investigate the genetic correlations with disease, explore functional enrichments, report on several genome-wide significant loci and the involvement of co-localized genes. On the basis of a single common variant, which has the largest effect on all properties of cfDNA, we investigate the causal mechanism and the clinical consequences for the implementation of NIPT, which likely translate to other forms of cfDNA screening.

## RESULTS

Keywords: NIPT, VeriSeq, fetal fraction, cell-free DNA, fragmentomics, DNASE1L3, rs35677470, PADI4, PANX1, DFFB

### GWAS of cfDNA identifies many genome-wide significant loci

GWAS was conducted for various measures related to cfDNA concentration and fragmentation. Concentration measures involved the total plasma cfDNA concentration, as well as the concentration stratified into fetal and mitochondrial origin (Figure 1). Fragmentation measures included size distribution, variable length nucleotide (and Purine/Pyrimidine) cleave site motifs, and the genomic distribution of cfDNA fragments (Figure 2). To assess the importance of these genetic factors in daily clinical practice, GWAS was also conducted to study the influence of common genetic variants on inconclusive NIPT results and the FF in a plasma sample (Table S1). We discovered many genome-wide significant loci and nearly all measures had significant heritability estimates (see Table 1).

**Figure 1:**
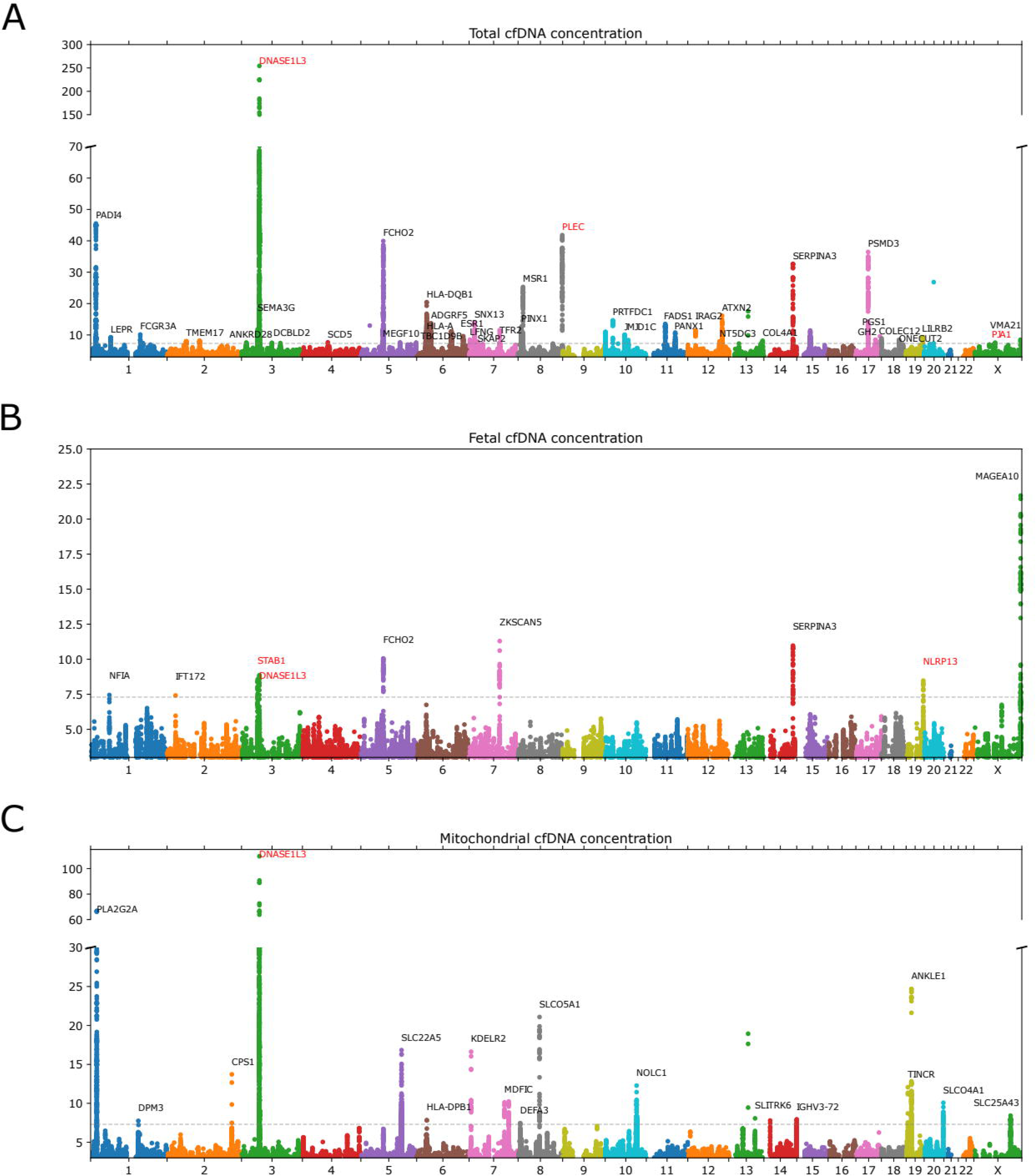
Manhattan plots for GWAS of stratified cfDNA concentration measures. Manhattan plots for (stratified) cfDNA concentration measures. For visualization purposes, gene annotations are limited to the strongest genome-wide significant associations to prevent overlapping labels. Loci were annotated with the nearest gene name up to a max distance of 100kb. Gene names are plotted in red when the variant intersected with the coding sequence of the gene. Variant associations with the concentrations of total (A), fetal (B) and mitochondrial (C) cfDNA. See also Table S1.

**Figure 2:**
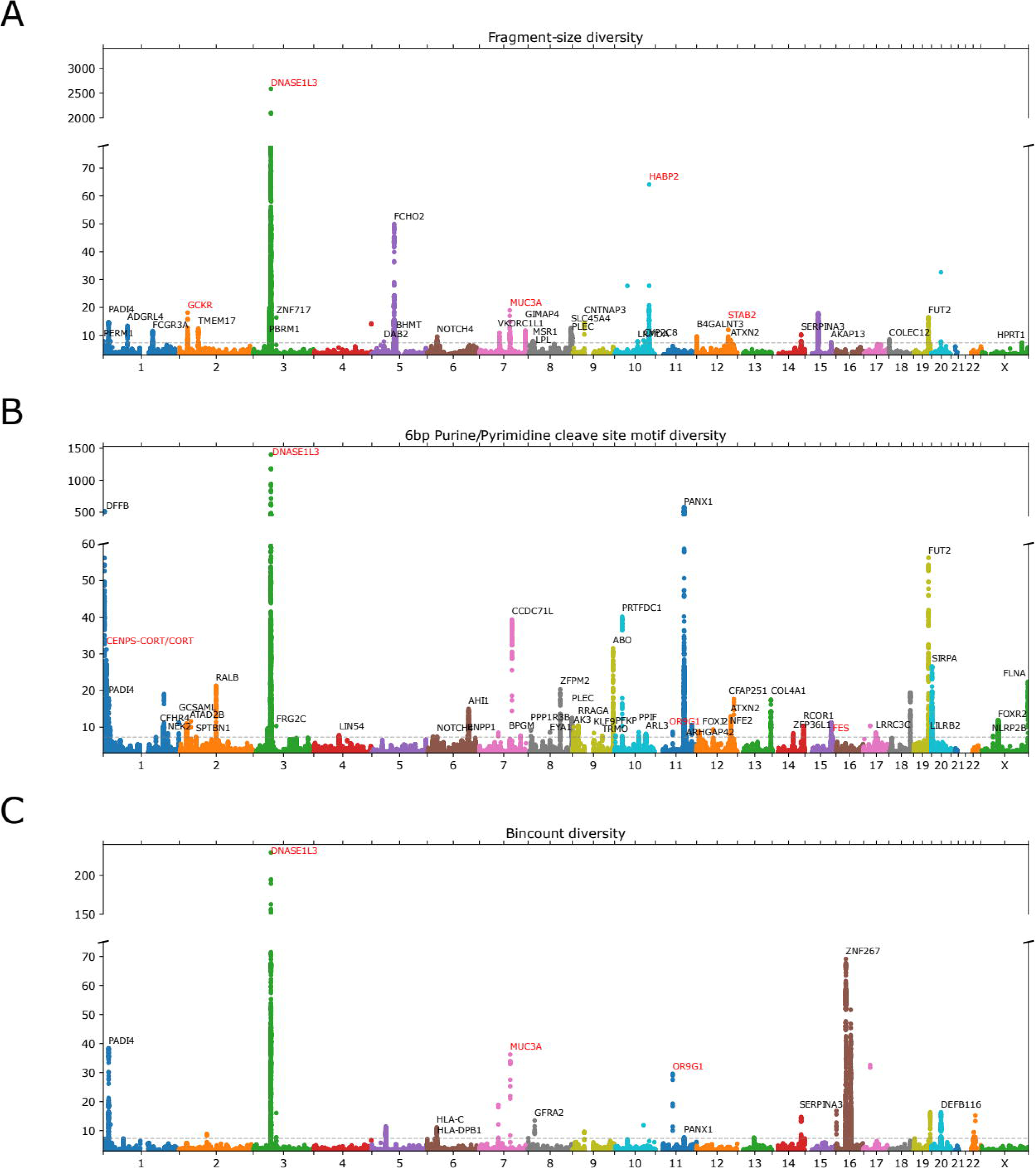
Manhattan plots for GWAS of fragmentomics measures. Manhattan plots for cfDNA fragmentation measures, using the VeriSeq FF measure as an additional covariate. For visualization purposes, gene annotations are limited to the strongest genome-wide significant associations to prevent overlapping labels. Loci were annotated with the nearest gene name up to a max distance of 100kb. Gene names are plotted in red when the variant intersected with the coding sequence of the gene. Variant associations with the diversity of cfDNA fragment sizes (A), 6bp Purine/Pyrimidine motifs (B) and the 50kbp bincounts across the genome (C). See also Table S1.

**TABLE 1:** Overview of all cfDNA GWAS results.

| Category | Phenotype | Description | N samples | Genome-wide significant loci <sup>a</sup> | LD score regression intercept <sup>b</sup> | LD score regression h2 <sup>c</sup> |
| --- | --- | --- | --- | --- | --- | --- |
| Calibration | Height | Maternal height | 140,962 | 424 | 1.1586 (0.0188) | 0.3792 (0.0197) |
|  | BMI | Maternal BMI (at time of blooddraw) | 140,955 | 140 | 1.1356 (0.0153) | 0.2938 (0.0125) |
| Concentration | Total cfDNA concentration | The total concentration of cfDNA in the sequencing library | 128,437 | 52 | 1.1231 (0.0111) | 0.1744 (0.0156) |
|  | Maternal cfDNA concentration (chrY) | The concentration of maternal derived cfDNA in the sequencing library | 66,048 | 14 | 1.0645 (0.0103) | 0.1978 (0.0199) |
|  | Fetal cfDNA concentration (chrY) | The concentration of fetal derived cfDNA in the sequencing library | 66,048 | 10 | 1.0258 (0.0092) | 0.1114 (0.0116) |
|  | Mitochondrial cfDNA concentration | The concentration of mitochondrial cfDNA in the sequencing library | 129,100 | 22 | 1.0351 (0.0106) | 0.0523 (0.0092) |
| Fragmentation | Fragment size diversity | Diversity of cfDNA fragment size distribution | 128,826 | 60 | 1.1075 (0.0113) | 0.1550 (0.0854) <sup>d</sup> |
| Genomic distribution | Bincount diversity | Diversity of cfDNA fragments in 50kbp bins across the genome | 129,123 | 26 | 1.2261 (0.0161) | 0.0245 (0.0151) <sup>d</sup> |
| Nucleotide cfDNA cleave site motifs | 2bp nucleotide cleave sites | Diversity of the 1bp nucleotide motifs up- and downstream of cfDNA fragment start and end sites | 129,123 | 9 | 1.0616 (0.0096) | -0.0005 (0.0045) <sup>d</sup> |
|  | 4bp nucleotide cleave sites | Diversity of the 2bp nucleotide motifs up- and downstream of cfDNA fragment start and end sites | 129,123 | 55 | 1.1075 (0.0119) | 0.1186 (0.0326) |
|  | 6bp nucleotide cleave sites | Diversity of the 3bp nucleotide motifs up- and downstream of cfDNA fragment start and end sites | 129,123 | 26 | 1.1065 (0.0122) | 0.0288 (0.0094) |
|  | 8bp nucleotide cleave sites | Diversity of the 4bp nucleotide motifs up- and downstream of cfDNA fragment start and end sites | 129,123 | 21 | 1.1066 (0.0126) | 0.0133 (0.0064) |
| Purine/Pyrimidine cleave site motifs | 2bp Purine/Pyrimidine cleave sites | Diversity of 1bp Purine/Pyrimidine motifs up- and downstream of cfDNA fragment start and end sites | 129,123 | 10 | 1.0357 (0.0097) | 0.0553 (0.0135) |
|  | 4bp Purine/Pyrimidine cleave sites | Diversity of 2bp Purine/Pyrimidine motifs up- and downstream of cfDNA fragment start and end sites | 129,123 | 57 | 1.1434 (0.0141) | 0.1884 (0.0576) |
|  | 6bp Purine/Pyrimidine cleave sites | Diversity of 3bp Purine/Pyrimidine motifs up- and downstream of cfDNA fragment start and end sites | 129,123 | 60 | 1.1341 (0.0132) | 0.1969 (0.0633) |
|  | 8bp Purine/Pyrimidine cleave sites | Diversity of 4bp Purine/Pyrimidine motifs up- and downstream of cfDNA fragment start and end sites | 129,123 | 26 | 1.1013 (0.0119) | 0.0277 (0.009) |
| NIPT Screening outcome | Type-1 | NIPT failure due to too little sequenced reads | 96,043 | 1 | 0.973 (0.0063) | 0.0013 (0.005) <sup>d</sup> |
|  | Type-2 | NIPT failure due to a too low fetal fraction (independent of BMI) | 96,043 | 3 | 0.9941 (0.006) | 0.0173 (0.0036) |
| Fetal fraction estimates | chrY FF | Fetal Fraction in male bearing pregnancies, derived from the Y chromosome | 37,868 | 23 | 1.1148 (0.0101) | 0.4813 (0.0256) |
|  | VeriSeq FF | Fetal Fraction in male bearing pregnancies, predicted from fragmentomics (proprietary) | 37,868 | 18 | 1.0936 (0.01) | 0.4397 (0.0403) |
|  | SeqFF | Fetal Fraction in male bearing pregnancies, predicted from fragmentomics (binned counts) | 37,868 | 16 | 1.0748 (0.0096) | 0.4205 (0.0317) |
|  | VeriSeq - chrY | Difference in FF estimates between chrY and VeriSeq | 37,868 | 8 | 1.0227 (0.0085) | 0.1343 (0.0417) |
|  | SeqFF - chrY | Difference in FF estimates between chrY and SeqFF | 37,868 | 5 | 1.0044 (0.0084) | 0.0528 (0.0154) |

<sup>a</sup> Number of independently associated loci above the genome-wide significance threshold ( $p < 5 \times 10^{-8}$ ), obtained from 'clumping' analysis.
<sup>b</sup> LD-score regression intercept (estimates the inflation in variant test statistics) after using for 20PCs as covariates, followed by standard deviation in parentheses.
<sup>c</sup> LD-score regression heritability estimates (the estimated amount of variation in the phenotype that can be explained by the imputed SNPs) followed by standard deviation in parentheses.
<sup>d</sup> Not significant at a 5% significance threshold.

It was found that the imputation quality, expressed as the average genotype probability, was negatively affected by increased fetal fractions (FF) and the number of sequenced reads in NIPT samples. NIPT-derived GWAS for two available maternal phenotypes: height and body mass index (BMI), resulted in high genetic correlations (rg) (rg=0.9617 and rg=0.8546 respectively) with publicly available GWAS summary statistics from the UK biobank (https://www.ukbiobank.ac.uk/) for the same traits, but based on dedicated SNP arrays.

To account for bias introduced by population structure, principal component analysis (PCA) was performed. Inspection of the first two PC’s indicated that although we observe large ancestral diversity in our dataset, about 87% of our samples overlapped with the EUR super population annotation of the 1000 genomes project^35^. For our cfDNA phenotypes, we observed minor, but significant effects amongst the first 17 PC’s. The strongest effect (Pearson r=0.092, p=3.97x10_-138_) was observed for total cfDNA concentrations along the first PC.

### R206C in DNASE1L3 affects all properties of cfDNA

Across all cfDNA GWAS (except the concentration of fetal cfDNA) the strongest and most significant effect was observed for a C to A missense variant (rs35677470) in *DNASE1L3*, which changes the Arginine at position 206 into a Cysteine (p.Arg206Cys, here referred to as R206C) and has a minor allele frequency of about 7% in the Dutch population. *DNASE1L3* encodes an endonuclease that is predominantly expressed by macrophages in the liver and is secreted into plasma. It has the unique ability to cleave lipid and protein bound DNA, and was previously found to be the main determinant of mono-nucleosome sized plasma cfDNA fragments.

R206C had the most significant effect on the size-diversity of circulating cfDNA fragments (beta=0.013; se=0.0001; p= 1x10^-2791^; Figure 2A). The R206C genotype was associated with a relative decrease in mono-nucleosome sized fragments and an increase in very short (50- 100bp) and di-nucleosomal fragments (Figure 3A). The R206C genotype has the most profound impact on cfDNA fragments over 400bp and thus beyond the size-range of our typical NIPT data.

**Figure 3:**
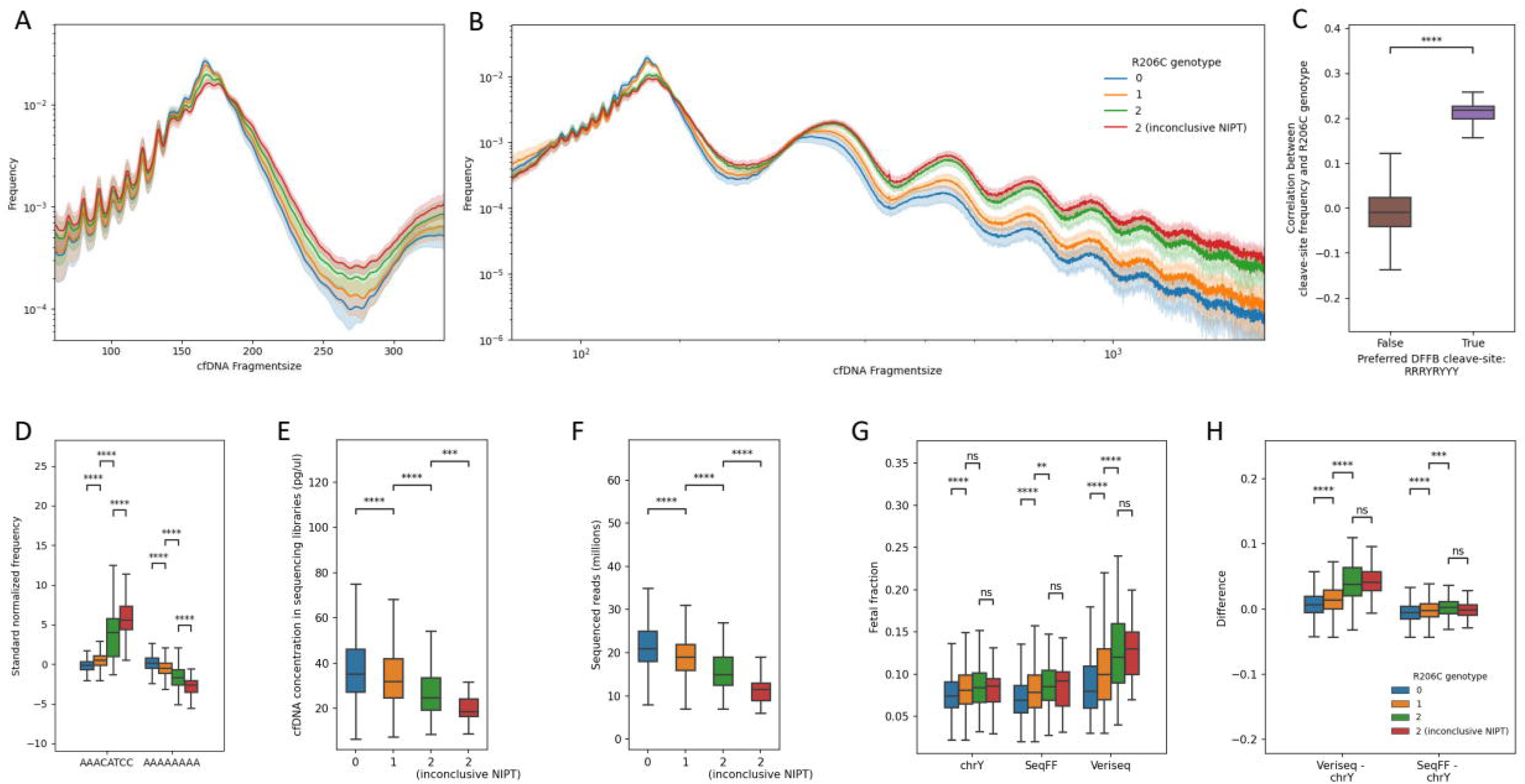
The effect of R206C on cfDNA. The effect of the imputed R206C genotype on the fragment size distribution of short cfDNA fragments (A). The effect of the ddPCR validated R206C genotype on the fragment size distribution of short and long (B) cfDNA fragments using long-read sequencing in the validation cohort of 55 samples. The colored line corresponds to the mean across each pool of samples, the shaded colored band corresponds to a 2 SD confidence interval (A and B). Boxplot showing the distribution of correlation coefficients between imputed R206C genotypes and nucleotide cleave-site motif frequencies that adhere to the RRRYRYYY motif (C). Boxplot showing the strongest positive and negative correlating 8bp cleave-site motifs (D). Boxplots showing the effect of the imputed R206C genotype on total cfDNA concentrations (E), uniquely aligned sequence (F), FF estimates (G), and differences in FF estimates with respect to the Y chromosome (H). See also Table S2.

Upon investigating all motifs up to 8bp (4bp up- and downstream) at cfDNA cleave sites, we found that R206C most strongly affected the diversity of 6bp Purine/Pyrimidine cleave-site motifs (beta=0.0005, se=5.55x-10^6^; p=1x10^1588^; Figure 2B). The frequency of almost all cleave site motifs was significantly altered by the R206C genotype (Table S2). Likely as a result of this, the genomic distribution of cfDNA fragments, as measured by the diversity of the number of reads across 50kbp bins throughout the genome, was also significantly altered by R206C (Figure 2C). We found the strongest correlations between the R206C genotype and the frequency of nucleotide cleave site motifs ‘ACGT’ and ‘AAACATCC’ (Figure 3D). The latter aligns with the ‘RRRYRYYY’ Purine/Pyrimidine motif, which was previously identified as the preferred cleavage site for DFFB^9^, whereas the former corresponds to the ‘RYRY’ central core of this motif. All other cleave-sites that conformed to this motif also positively correlated with the R206C genotype (Figure 3C). Among the 100 strongest correlating motifs, there were 34 other ‘RRRYRYYY’ Purine/Pyrimidine motifs (Table S2). Amongst the 100 strongest correlating motifs, the ‘AAAAAAAA’ motif was the only motif for which the frequency negatively correlated with the R206C genotype (Figure 3D and Table S2).

We found that R206C caused a significant reduction (beta=0.116; se=0.003; p= 1.29x10^-2^^63^) in total plasma cfDNA concentrations (Figure 3E), which despite equimolar pooling of samples throughout sequencing runs, also resulted in a significant decrease in the number of sequenced cfDNA fragments (Figure 3F).

### R206C increases risk of inconclusive NIPT

As we observed large effects of R206C on all cfDNA properties, we wondered if R206C and other variants affected our NIPT screening results during routine clinical practice. We performed case/control GWAS to investigate the effect of common SNPs on inconclusive screening outcomes. In practice, at our lab, the causes of inconclusive NIPT (after repeated sequencing experiments on the same blooddraw) are either related to impaired sequencing yield (e.g. less than 5 million uniquely aligned fragments), here defined as type-1, or very low FF (<1%), defined as type-2. Type-1 inconclusive NIPT at our facility has a prevalence of ∼0.37%, while type-2 inconclusive NIPT has a prevalence of ∼2.25%.

GWAS for type-1 inconclusive NIPT resulted in only one genome-wide significant locus: R206C (OR=7.03; log(OR)=0.85; log(OR)_se=0.08; p=8.3x10^-127^). On the basis of our imputed genotypes, homozygosity for R206C increases the odds of type-1 inconclusive NIPT results ∼14-fold (Table S1). Our GWAS for type-2 inconclusive results identified 3 loci (Table S1), also including the R206C variant in *DNASE1L3*. However, in this case R206C had a ‘protective’ effect as it decreased the odds of NIPT failure due to a too low FF (OR=0.54; log(OR)=-0.27; log(OR)_se=0.07; p=6.37x10^-19^).

### R206C increases fetal fraction

The sensitivity of NIPT is largely dependent on the fraction of sequenced fetal DNA, which is derived from the placenta and varies significantly between pregnant women. For this reason, the FF is an essential parameter for NIPT. In male-bearing pregnancies, the FF can be derived from the sex-chromosomes. To also obtain FF estimates in female-bearing pregnancies, fragmentomics-based predictors, such as the VeriSeq (Illumina, San Diego, USA) FF solution^36^, have been developed. The SeqFF^37^ predictor makes use of the differential genomic^37^ distribution, while the VeriSeq predictor also makes use of the differential size^38^ of maternal and fetal cfDNA.

Our findings with respect to the observed ‘protective’ effect of R206C on type-2 inconclusive NIPT results had two possible explanations. Either FF was truly increased in NIPT samples with this genotype, indicating a biological effect, or it was the result of the perturbed fragmentomics affecting the FF predictors, or a combination of both.

To address this question, we selected only male-bearing pregnancies, where we could use the Y chromosome (chrY) as a fragmentomics-free alternative to the SeqFF and VeriSeq predictor^36^ to study FF. We performed GWAS on all three FF measures and found that results were highly correlated (LD-score regression based genetic correlation: chrY/ VeriSeq=0.9733; chrY/SeqFF=0.9766; VeriSeq/SeqFF=0.9734, Table S1). For all FF measures the most significant locus was R206C. While the direction of effect was the same, the estimated effect sizes differed significantly. The most significant effect of R206C was observed for the VeriSeq predictor (Beta=0.016, se=0.00055, p=2.84x10^-190^), followed by SeqFF (Beta=0.010, se=0.00036, p=1.84x10^-176^). The effect of R206C on the Y-chromosome derived FF was smaller (Beta=0.007, se=0.00033; p=4.14x10^-110^), but still highly significant.

### Fetal fraction increases as a result of decrease in maternal cfDNA

We evaluated whether the R206C variant affected the fetal fraction by increasing the amount of fetal DNA, or by decreasing the amount of maternal DNA, or both. To address this question, we stratified the total cfDNA concentration into fetal and maternal concentrations by multiplying it with our FF estimates. When we used these measures as GWAS targets, we observed a genome-wide significant, cfDNA-decreasing effect of the R206C genotype on both. However, the effect on the maternal cfDNA concentration was much stronger than the effect on fetal cfDNA, explaining the increased FF. Interestingly, the fetal cfDNA concentration GWAS did result in several other more significant loci (Figure 1B). To validate this approach, we stratified cfDNA concentrations based on the mitochondrial fraction of cfDNA (which also varies and can easily be derived for all cfDNA samples). Using this GWAS we identified many genome-wide significant loci (Figure 1C), which functionally implicated mitochondrial function and were highly distinct from the loci identified using our total cfDNA concentration GWAS, proving the validity of our approach.

### Altered fragmentomics contributes to the differences in fetal fraction prediction

Despite the fact that all three FF measures were highly correlated (Pearson correlation with chrY/ VeriSeq =0.87; chrY/SeqFF=0.83; VeriSeq /SeqFF=0.84) and R206C seemed to exhibit a true biological effect on FF, we observed significantly different effect sizes, while all studies were performed on the exact same dataset. As a result, we wondered whether the altered fragmentomics, as a result of common genetic variants, contributed to these differences. To investigate this, we calculated the difference between the Y chromosome derived FF and the predictions by both VeriSeq and SeqFF, and used these as GWAS targets (Table S1). We identified 6 and 4 genome-wide significant hits and obtained heritability estimates of ∼11% for the difference between Y chromosome derived FF and VeriSeq, and ∼4% for the difference between Y chromosome derived FF and SeqFF. Indicating that a significant fraction of the difference between the methods is not the result of random error, but instead is attributable to genetic variation between the samples. Again, the strongest effect was observed for the R206C variant on VeriSeq FF predictions (Figure 3G and 3H). For both methods, the R206C variant resulted in an overestimation of the FF compared to the chromosome Y based estimate. In case of homozygosity for the R206C variant, VeriSeq on average overestimated the FF with approximately 4.2%.

### Targeted genotyping strengthens the association with inconclusive NIPT

We imputed the R206C genotype with high confidence (INFO score: 0.90269). Nonetheless, we validated the imputed R206C genotype using left-over plasma from a total of 63 NIPT samples and used digital droplet PCR (ddPCR) for direct genotyping (Table S2). We selected 26 samples for which we imputed a homozygous R206C allele (11 of which resulted in type-1 inconclusive NIPT results and 15 with conclusive NIPT results and no detected trisomies), 15 samples for which we imputed a heterozygous allele and 22 samples for which we imputed the wild type allele (all with conclusive NIPT results). Across the four groups samples were selected at random. On average we obtained 220 (sd=117) allele counts (droplets that were either positive on channel 1 or channel 2) per sample, from which we inferred the most likely maternal genotypes based on a simple Bayesian model (see Method details). We found that for 57 out of 63 (91%) the imputed genotypes were identical to the ddPCR inferred maternal genotypes. In 6 samples we found that ddPCR indicated a heterozygous allele while the homozygous allele was imputed. These were all samples from the group of 15 for which sufficient sequencing data was generated, and normal conclusive NIPT results were obtained. Suggesting that the combined evidence of inconclusive NIPT results and an imputed homozygous R206C allele provided additional evidence for the true R206C genotype. We indeed found that the effect of imputed homozygosity for R206C in combination with inconclusive NIPT results was stronger than the imputed R206C genotype alone (see Figure 3A,D,E,F,G,H).

We also found that in this smaller validation set the R206C ddPCR genotype was also significantly associated with a lower cfDNA concentration measure, both before (Mann-Whitney-Wilcoxon test two-sided test, before p=7.822e-03) and after library preparation (p=2.471e-03).

### Long-read sequencing shows increase in very large cfDNA fragments

Besides a lower cfDNA concentration, we also found that R206C overall caused a small increase in the size of sequenced cfDNA fragments. We therefore hypothesized that the reduced enzymatic activity of DNASE1L3 would lead to longer cfDNA fragments in plasma, which we cannot observe with NIPT as they are outside the size-range of Illumina sequencing technology (>400bp). To investigate this, we used plasma DNA from 55 of the 63 samples for which the R206C genotype was previously validated using ddPCR and material was still available. This resulted in 20 homozygous R206C, 15 heterozygous and 20 homozygous wildtype samples for the R206C allele, which were simultaneously sequenced on a single PromethION (Oxford Nanopore, London, UK) 2 flowcell.

On average we obtained 1.66 million reads per sample. By pooling samples we observed an additive effect of the R206C genotype on the frequency of very long cfDNA molecules (Figure 3B). Despite the fact that all genotypes were validated using ddPCR and incorrectly imputed genotypes should therefore not play a role, we still found that the effect was stronger in the group with inconclusive NIPT results.

The reduction in short mono-nucleosome fragments, which we observed previously from the NIPT short-read data (Figure 3A) was also apparent in the long-read sequencing data (Figure 3B). One sample, which was homozygous for the R206C allele and had conclusive NIPT results, exhibited a highly distinct fragment-size distribution and was therefore excluded from our analyses. Despite equimolar pooling of cfDNA quantities, we still observed that the R206C allele significantly decreased the number of sequenced cfDNA molecules.

### Other genome-wide significant variants implicate nucleases, phagocytes and NETs

Our GWAS also identified many other genome-wide significant loci (Table S1). Beyond *DNASE1L3*, the classic model of cfDNA biology features two other genes: *DFFB* and *DNASE1*. While we did not observe an effect on cfDNA from common variants near *DNASE1*, we did observe highly significant associations between intronic variants (lead SNP rs34141819) in DFFB and the motif-diversity of cfDNA cleave sites. Beyond its effect on cleave-site motifs, this variant did not have a significant effect on the size-distribution or concentration of cfDNA (Figures 1 and 2), which is in line with previous work in *DFFB*-deficient mice^5^. The motif- diversity GWAS also identified a third highly significant association with variants in the Pannexin-1 (*PANX1*) gene (rs4753126 and rs1138800), of which rs1138800 encodes for a gain-of-function variant (Panx1-400C) and was previously associated with enhanced platelet reactivity^39^. The *PANX1* gene is a widely expressed cell-membrane channel that during apoptotic cell death is involved in the formation of membrane protrusions and the release of ATP into the extracellular space, which acts as a ‘find-me’ signal for macrophages. The same variant in *PANX1* was also significantly associated with all other concentration and fragmentation properties of cfDNA.

Variants were also identified in or near other genes that were previously associated with cfDNA biology, such as Complement Factor H (*CFH*) and Hyaluron Binding Protein 2 (*HABP2*). *HABP2*, also known as Factor-7 activating protease (*FSAP*), is involved in the release of nucleosomes from late apoptotic cells^7^. In this gene we observed a missense variant with a minor allele frequency of 3%, called the Marburg 1 polymorphism^40^, which also has a relatively large effect on the fragmentation patterns of cfDNA. Resulting in a relative increase in short (<166bp), but a decrease in both mono- and multi-nucleosomal fragments.

Beyond digestion by blood-circulating nucleases, the clearance of apoptotic bodies and cfDNA is further regulated by their uptake by macrophages in the liver and spleen^15^. We detect multiple genome-wide significant loci across multiple cfDNA traits, near macrophage scavenger receptors *MSR1, COLEC12, STAB1* and *STAB2*, which can bind to exposed ligands on the surface of apoptotic cells and play an important role in the recognition and uptake of cellular debris from apoptotic cells. We also detect a block of variants in high linkage, which span the genes Transportin-1 (*TNPO1*) and the Fer/CIP4 domain-only protein 2 (*FCHO2*), *FCHO2* is involved in clathrin-mediated endocytosis and is predominantly expressed in macrophages and Kupffer cells.

Several loci seem to link cfDNA to neutrophils and the NETosis cell-death pathway. Protein-arginine deiminase type-4 (*PAD4*) is involved in the citrullination of histones, which plays an important role in the decondensation of chromatin during NETosis and the formation of neutrophil extracellular traps (NETs). The *PAD4* locus mainly affects the concentration of cfDNA. Another locus that mostly affect cfDNA concentrations is upstream of *SERPINA3* and a cluster of other serine protease inhibitor genes. *SERPINA3* is an inhibitor of chymotrypsin-like proteases, such as Cathepsin G and neutrophil elastase, which are expressed in neutrophils, are abundantly present in NETs and have roles in both the formation and degradation of NETs. Our GWAS also detects a block of variants in high-LD which spans the genes: GSDMA/PSDM3/CSF3/MED24, which affects both the concentration and fragmentation patterns of cfDNA. This locus has previously been associated to eosinophil counts and asthma. Within this locus Gasdermin-A (*GSDMA*) is a plasma membrane channel related to pyroptosis, a regulated inflammatory cell-death mechanism, while the Granulocyte colony-stimulating factor (CSF3), encodes a protein that stimulates the bone marrow to produce granulocytes such as neutrophils and eosinophils.

Furthermore, we find a strong association between a common missense variant (rs6992333) in Plectin (*PLEC*) and multiple cfDNA traits. Plectin is a widely expressed protein which links the three main components of the cytoskeleton as well as the plasma membrane. Although the specific role of PLEC in cfDNA biology is not fully understood, Plectin was found to be a substrate for caspases, contributing to the cytoskeletal rearrangements during apoptosis^41^.

### Variants that specifically affect circulating placental DNA

We identified multiple variants that were specifically associated with the concentration of fetal derived cfDNA fragments. Amongst these were variants in NOD-like receptor family pyrin domain containing 13 (*NLRP13*) and Stabilin-1 (*STAB1*). The missense variant in NLRP13 was previously associated with offspring birth weight^42^, while *STAB1* is a receptor expressed on placental macrophages and has a function in clearing apoptotic cells at the feto-maternal interface^43^. *STAB1* double knockout-mice were previously found to suffer from defects in placental development, which was explained by the role of this gene in remodeling the spiral arteries during placental development^44^.

### Genes that affect the concentration of mitochondrial cfDNA

As mentioned previously, to validate the approach to distinguish total amounts of fetal and maternal cfDNA in plasma samples, we performed a similar analysis to investigate variants that affected the amount of mitochondrial cfDNA (mtcfDNA). The analysis resulted in several genome-wide significant loci. These loci are interesting as mtcfDNA is a highly potent inflammatory trigger^45^. Besides the association of R206C in *DNASE1L3*, which caused an increase in the amount of mtcfDNA in plasma, the second most significant locus was observed near the membrane associated phospholipase A2 (*PLA2GA*). *PLA2GA* plays an important role in regulating vascular inflammation. The lead-variant in our GWAS is in high-linkage with rs4744, a protein truncating mutation, which was previously associated with variable serum levels of *PLA2GA* and c-reactive protein. The third most significant locus was found near Ankyrin Repeat and LEM Domain Containing 1 (*ANKLE1*). *ANKLE1* is an endonuclease^46^, colocalizes to mitochondria and was recently found to cleave mitochondrial DNA during erythropoiesis^47^.

### Heritability estimates and genetic correlations

Besides exploring the genes in loci with a relatively large effect, we also explored the joined effect of thousands of loci with relatively small effects on cell-free DNA. We used LD-score regression^48^ to calculate the common SNP heritability (h^2^) of all cfDNA traits and found significant heritability estimates for almost all (Table 1). The highest heritability estimates were observed for FF, the size and 6bp Purine/Pyrimidine cleave-site motif diversity of cfDNA fragments. For our GWAS studies of cfDNA, we observed multiple significant genetic correlations with the risk of different systemic autoimmune (such as SLE, Rheumatoid Arthritis and Primary biliary cholangitis) and cardiovascular diseases (such as Stroke and Coronary Artery disease), see Table S3.

Finally, we used the stratified or partitioned heritability approach of LD-score regression to determine heritability estimates for different functional genome annotations^49^. We used the cell/tissue type specific genome annotations from the baseline model v2.2^49^. When we investigated different cfDNA concentration GWAS results, we observed moderate, but significant enrichment for heritability in regions that are expressed (or in specific chromatin-states) in Phagocytes, Liver, Adipose tissue, and (adipose tissue) Macrophages (Table S3), all known to be involved in cfDNA biology.

## DISCUSSION

A considerable amount of variation in cfDNA properties can be observed within the general population. The factors that influence these properties are incompletely understood. Our study shows that a significant proportion of this variation can be attributed to genetic variation in the form of common SNPs.

### DNASE1L3 R206C: clinical implications

The largest contribution of a single variant is observed for R206C, a missense variant in DNASE1L3. In the Dutch population this variant has an allele frequency of ∼7%. This has consequences for clinical use of NIPT, as this variant causes an increase in FF. Furthermore, the differential fragmentation of cfDNA as a result of this variant causes predictors to overestimate the FF. To illustrate, in homozygous carriers of the R206C variant, VeriSeq on average predicts the FF to be ∼4.2% higher than the FF that can be inferred from the Y chromosome. The reason the VeriSeq FF predictor is more affected than the SeqFF predictor may results from the fact that only VeriSeq uses the size of cfDNA fragments to predict FF, and the R206C variant most strongly affects this property of cfDNA. VeriSeq likely uses the fact that placenta-derived cfDNA fragments are slightly shorter than cfDNA fragments derived from other cells^38^. While we observe a strong increase in long cfDNA fragments, we also note that R206C seems to cause a relative increase in short fragments (<140bp, see Figure 3A). Other applications of prenatal cfDNA screening, which also employ this characteristic to distinguish between maternal and placental origins, may therefore be affected by R206C as well^50–53^.

The increased size of circulating cfDNA fragments, in combination with a reduced cfDNA concentration, decreases the number of sequenced fragments in R206C carriers. cfDNA sequencing of homozygous carriers of this variant frequently resulted in less than 5 million sequenced cfDNA fragments (10 million reads), less than half of what is generally obtained, despite equimolar pooling of samples. At our facility, homozygosity for this variant accounted for a ∼14x increased odds of receiving type-1 inconclusive NIPT results. We conclude that this is a conservative estimate as this risk calculation was based on imputed genotypes, which upon ddPCR based validation, were found to contain a considerable amount of genotyping errors. However, when these errors are accounted for, we find that there still is significant variability in the size-distribution when comparing samples with and without conclusive NIPT results and the same genotype. This either suggests a polygenic effect, for which we did not find any obvious evidence, or potentially a non-genetic effect on DNASE1L3 function. In this, we note that the impaired function of DNASE1L3 may also be autoantibody-mediated^32,54^. The impaired function of DNASE1L3 may also provide a causal link for the observation by others that autoimmune diseases are overrepresented in inconclusive NIPT samples^55^ and that immune-mediated diseases can be predicted from NIPT data^56^.

So far, at our facility we found evidence for one case of trisomy 18 with inconclusive NIPT results as a consequence of the mother being homozygous for the R206C variant. Generally, caution is warranted in interpreting wgs-based NIPT results from R206C carriers, as in the case of VeriSeq (and likely also other implementations) the overestimated fetal fraction is used in the statistical test to determine a fetal genomic aberrations. This in combination with the reduced sequencing yield, suggests an increased odds for false negative screening results. To counter this, efforts should aim to further improve both sequencing yields from low cfDNA input concentrations and WGS-based FF predictors. Either by explicitly accounting for the R206C genotype, or to employ a genome-wide polygenic scoring algorithm to boost predictions.

Although much of the observed increase in FF amongst R206C carriers seems to be attributable to difficulties of predictors in modelling the differential cfDNA fragmentation patterns, we do observe a smaller, but highly significant effect on FF through Y-chromosome based estimates. We pose that this observation may underlie a true biological effect, which is in line with recent work that found that by impairing the clearance of cfDNA by means of antibodies or liposomes, could significantly improve the sensitivity of circulating tumor DNA tests^57^. Alternatively, it was also found that in DNASE1L3^-/-^ mice, pregnancy with DNASE1L3^+/-^ fetuses, could partially reverse the aberrant cfDNA profile^58^. Also in humans, differences between the maternal and fetal genotype may therefore play a role. Unfortunately, these samples typically have very low cfDNA input concentrations, which makes it hard, even with targeted approaches, to study the fetal genome from cfDNA.

These findings are also relevant for other forms of non-invasive screening, such as liquid biopsies^59^. Also those that depend on methylation signatures of cfDNA^60^, as it was previously shown that DNASE1L3 prefers cleavage at methylated CpGs and complete DNASE1L3 deficiency resulted in altered cfDNA methylation profiles^61^.

### DNASE1L3 R206C: lowered cfDNA concentration

It was previously found that R206C impairs the secretion of DNASE1L3, and thereby limits the enzymatic activity of DNASE1L3 in plasma^29^. We find that this impaired secretion results in decreased cfDNA concentrations of our plasma DNA sequencing libraries. From our validation tests we find that, also in R206C carriers, the cfDNA concentration before and after library preparation are highly correlated, suggesting that the observation is not caused by the library preparation protocol. We therefore propose that the impaired secretion of DNASE1L3 causes DNA fragments to remain in incompletely cleared cellular debris (e.g. NETs, extracellular vesicles, exosomes, micro vesicles, apoptotic bodies or immune complexes), which are either disposed of by alternate biological pathways or during the centrifugation stage of plasma extraction.

### Contribution of NETs to cfDNA

Most cfDNA molecules in plasma have been shown to derive from apoptotic hematopoietic cells. However, under specific pathological conditions, alternative cell-death mechanisms such as NETosis, are increasingly recognized as contributors to the pool of plasma cfDNA (e.g. autoimmune disease, thrombosis, infections etc.). In these studies, cfDNA concentrations are often used as a proxy to study NET formation. Our study amongst the general population of healthy pregnant women detects a locus near the PADI4 gene which plays an important role in the initiation of NET formation, which suggests that NETs also contribute to the cfDNA pool under physiological conditions.

### Nuclease genes and cfDNA: two nucleases and PANX1

GWAS on cleave-site motifs at the extremes of cfDNA molecules revealed three highly significant loci in the genome, including *DFFB* and *DNASE1L3*. The third locus resides within the *PANX1* genes, which encodes a membrane channel through which ATP is released during cell-death. As *PANX1* is widely expressed in different cell types and has not been characterized as an endonuclease itself, we pose that *PANX1* may mediate the amount of extracellular DNA cleavage by circulating nucleases. Interestingly, we also find that as a result of R206C, the impaired *DNASE1L3* activity results in an increased frequency of DFFB-preferred cleave-sites motifs. Suggesting that the relative frequency of ‘RRRYRYYY’ cleave-sites motifs in cfDNA can be employed as a biomarker for *DNASE1L3* activity.

### Limitations, Open challenges and future work

Our work demonstrates the power of using widely available NIPT sequencing data to study the genetics behind cfDNA, a complex biological phenotype. Provided that phenotypical data is available, imputed variants from NIPT data offers and excellent opportunity to also study other complex traits and diseases. However, several challenges remain. For example, imputation accuracy should be further improved, specifically for variants with lower allele- frequencies. This is something that may be addressed by expanding reference datasets, but more importantly for the application of NIPT data is that current imputation models do not account for the paternal haplotype that is present in NIPT data. There is great potential for imputation methods that are not only faster and able to handle larger reference sets, but also specifically tailored to impute variants from NIPT data which is more complex, but also richer than typical whole-blood sequencing data.

Our study investigated the genetic architecture of cfDNA using a population of Dutch pregnant women. While future research should address other populations of individuals and pathological and non-pregnancy conditions, our work provides a basis for further improvements in non-invasive testing.

## Data Availability

All data produced in the present study are available upon reasonable request to the authors

## ACKNOWLEDGEMENTS

We’d like to acknowledge all pregnant women that consented for the use of their data, and all members of the NIPT laboratory in the Amsterdam UMC for generating the sequencing data. We’d also like to thank the Core Facility Genomics. In particular we’d like to thank the following people: Lars Nooi, Kavish Kohabir, Wouter Segerink, Roxane Boyer and Daoud Sie. This study uses data that was generated as part of the Dutch TRIDENT-2 study, for which we’d like to acknowledge all members of the Dutch NIPT consortium. The TRIDENT-2 study was funded by a grant from the Netherlands Organization for Health Research and Development (ZonMw, No. 543002001).

## AUTHOR CONTRIBUTIONS

Conceptualization, J.L. and M.N.; Methodology, J.L.; Validation, J.L. and M.N.; Formal Analysis, J.L.; Investigation, J.L.; Resources, E.S.; Data Curation, J.L. and E.S.; Writing – Original Draft, J.L.; Writing – Review & Editing, M.N. and E.S.; Visualization, J.L.; Supervision, E.S.; Project Administration; J.L. and E.S.; Funding Acquisition, E.S.

## STAR * Methods

### RESOURCE AVAILABILITY

#### Lead contact

Further information and requests concerning resources should be directed to Jasper Linthorst.

#### Materials availability

This study did not generate new unique reagents.

#### Data and code availability

- Unfiltered GWAS summary statistics are made available through locuszoom:

- Fragment size diversity: https://my.locuszoom.org/gwas/443494/?token=132c7e351ba84717a3f003299cd8d1f3
- Cleave-site motif diversity: https://my.locuszoom.org/gwas/416825/?token=50123122e9b347909414a1aa2ae032dd
- Bincount diversity: https://my.locuszoom.org/gwas/694315/?token=feef59a3e58f41ff8c14f5336ed808fc
- Total cfDNA concentration: https://my.locuszoom.org/gwas/310546/?token=5f45fdc6ab774947872221cb25fdeb4f
- Fetal cfDNA concentration: https://my.locuszoom.org/gwas/964912/?token=a8891852aa1b4143b242a25b1af521a0
- Mitochondrial cfDNA concentration: https://my.locuszoom.org/gwas/326281/?token=399f7ffd26504c3faf965a7080477173
- Individual imputation and sequencing data will not be made publicly available.
- All other data is available through the Supplementary Tables or upon request to the lead contact.

## EXPERIMENTAL MODEL AND STUDY PARTICIPANT DETAILS

We used low-pass whole-genome sequencing data from about 140,000 Dutch pregnant women from the general obstetric population collected in the Amsterdam UMC (region Amsterdam and the northern provinces of The Netherlands) between 2018 and 2021 as part of the TRIDENT-2 study^33^. Written informed consent was obtained from all participating women. Approval for the study was granted by the Dutch Ministry of Health, Welfare, and Sport (license 1017420-153371-PG) and the Medical Ethical Committee of VU University Medical Center Amsterdam (No. 2017.165). Samples in which trisomies were detected, and women that did not consent to the use of their data for scientific research were excluded from the analyses.

## METHOD DETAILS

### Sample collection

Blood draws were scheduled at or after 11+0 weeks of gestation. Sampling was performed at different service locations across the Netherlands. Blood was drawn in two 10 mL cfDNA BCT CE tubes (Streck) and shipped at room temperature by courier or regular mail in specific transport containers. Time between blood collection and plasma isolation was five days at most.

### Short-read cfDNA sequencing

Plasma isolation, cfDNA extraction, library preparation, quantification and sequencing were performed in accordance with the VeriSeq NIPT method (Illumina, San Diego, USA). Briefly, upon isolation of cfDNA from 1ml of plasma, samples were paired-end sequenced using 36bp on an Illumina NextSeq500. Raw base call files were de-multiplexed, and adapters were trimmed using bcl2fastq (2.17.1.14). All sequencing data was produced across several NextSeq500 sequencers at the Amsterdam UMC.

### Long-read cfDNA sequencing

DNA was extracted from 3 to 4ml left-over plasma using QIAsymphony DSP Circulating DNA Kit (protocol: circDNA3 CR22161 ID2686). The sequencing library preparation protocol SQK- NBD114.96 was used in combination with the NEB Next Companion Module (E7180S). Several modifications were made to the original protocol.

For DNA end repair in step 8, both the 20°C and 65°C steps were extended to 30 minutes each. During native barcode ligation in step 6, all DNA (15µL) was added, necessitating adjustments to all volumes: 5µL barcode and 20µL Blunt/TA Ligase Master Mix (total 40µL). Then in step 8 incubation was carried out for 4.5 hours at 20°C followed by overnight incubation at 4°C instead of the usual 20-minute incubation. In step 9, 4µL EDTA (clear cap) was added instead of 1µL. In step 10, pooled samples (∼2.1mL) were placed in a single 5mL tube. In step 12 the reaction was then split into two 2mL tubes for a 0.8x (800µL Ampure per tube) cleanup, instead of the standard 0.4x. Finally, during step 20, an eluate volume of 62µL was obtained in one tube, double the usual volume, necessitating the doubling of reagents for the ligation of the NA adapter. For adapter ligation and clean-up, the duration of step 7 was extended to 90 minutes, and 40µL was used instead of 20µL in step 9. Sequencing was performed on the ONT Promethion 2 Solo with a FLO-PRO114M Flowcell (version R10.4.1). Sequencing was performed for approximately 24 hours before the flow cell was washed with the Flow Cell Wash Kit (EXP-WSH004) and reloaded with fresh samples. Sequencing was stopped after 72 hours. Basecalling was performed using Dorado (version 7.2.13) with the dna_r10.4.1_e8.2_400bps_sup (v4.3.0) model.

### Processing sequencing data

In case multiple sequencing datasets were available for the same woman, the dataset which resulted in most sequencing data was included. Sequencing data was aligned to the human reference genome GRCh38 (excluding alt and unplaced contigs) using BWA-mem. Duplicate reads were filtered using Samblaster. Samples for which less than 5 million reads could be uniquely aligned, were excluded from the analyses. On average we sequenced 22 million, 36bp paired-end reads per sample.

### Imputation of genetic variants

Approximately 8 million SNPs with a MAF greater than 1% were selected from the Human Reference Genome Consortium dataset^62^ using bcftools^63^. These variants were transferred to GRCh38 coordinates using CrossMap^64^ and subsequently assigned to 25MB blocks across the human genome. All samples were simultaneously imputed for each of these blocks using QUILT^65^. QUILT version 0.1.9 was run with the following parameters: nGen=100 and buffer=100.000. The Human Reference Genome Consortium dataset^62^ was used as a reference.

### GWAS analyses

GWAS analyses and PCA were performed using Plink2^66^. The first 20 PC’s were used as covariates for all GWAS. Samples were not filtered out on the basis of the inferred ancestry. Additionally, BMI is known to affect FF^67^, and FF in turn can be predicted from fragmentomic features^37^. Where applicable, we therefore used BMI and FF (calculated by VeriSeq) as covariates to study all other traits. For LD-score regression, the resulting summary statistics were filtered for MAF>0.05 and INFO>0.6. Additionally, for visualization (in Figures 1 and 2) we filtered sites for which the allele frequency estimated from the overall ‘sequencing pile- up’ differed more than 0.2 from the imputed allele frequency. All cfDNA phenotypes were filtered by removing outliers that were more than 5 standard deviations away from the mean value. In case multiple concentration measurements were available for a plasma sample we used the mean of these values. To account for the right-tailed distribution of cfDNA concentration measures, data was log-transformed before GWAS analysis. All reported effect sizes and p-values are reported with respect to the log-transformed data. We used the clumping procedure as implemented in Plink (version 1.9) to report independently associated loci. Despite the use of strict clumping parameters (clump-p1=5e-8, clump-p2=5e-5, clump-kb=1000, clump-r2=0.01), the effect of the R206C variant on many cfDNA traits was so strong that variants up to 5Mbp up- and downstream were ignored in our search for other independent associations. As inconclusive NIPT results at our department were handled differently from the start of 2021, only samples up to 2021 were included in the related GWAS.

### Fragmentomics measures

We calculated fragmentomics features from the sequencing data itself. To derive cfDNA fragment-size distributions, we used the insert-size of paired-end sequenced fragments, computed by samtools^68^ stats in the range of 50 to 600bp. The genomic distribution of cfDNA fragments was calculated using samtools and bedtools^69^ (subcommand coverage). Cleave-site motifs were obtained using pysam^70^ (https://github.com/pysam-developers/pysam).

### Diversity measures: entropy calculation

Calculated fragmentomics measures result in a distribution of values for each sample. To perform GWAS, we calculated measures of diversity^71^ (normalized Shannon entropy) for each of these distributions. Briefly, we compute this measure as follows:

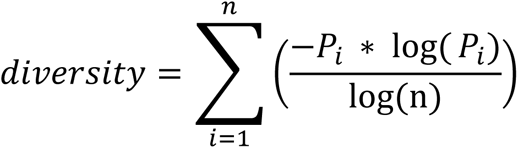

Where *n* is the number of measures in the distribution and *P_i_* is the frequency of the *i* th measure.

### Fetal fraction calculations

Fetal fractions were computed in three different ways: VeriSeq, SeqFF and based on the Y-chromosome. The VeriSeq FF method is proprietary and details about the method are undisclosed^36^. The SeqFF model was built on a dataset that was aligned to the GRCh37 assembly, it was therefore only run on a subset of our data for which alignments to this assembly were already available. To infer FF from the Y-chromosome we adopted the method referred to as DEFRAG_b_ in ^72^. Briefly, the median read count in 1MB bins across the Y-chromosome (excluding PAR regions and bins 0-2Mb, 9-14Mb, 20-21Mb and 25-59Mb) is divided over the median count in 1MB bins across the autosomes (excluding the chromosomes 13,18 and 21), and multiplied by 2. To select pregnancies where the fetus was male, we selected the top 51.22% of the samples with the largest Y-chromosome based FF estimate. This selection is in line with the 105 to 100 male-to-female birth ratio in The Netherlands and corresponded to a minimal chrY-based FF estimate of 2.2%.

### Targeted R206C genotyping

Bio-rad digital droplet digital PCR (ddPCR) was used to perform targeted genotyping of the R206C SNP. Probes were designed through the online Bio-Rad Mutation Detection assay. DNA extraction was performed using the QIAsymphony DSP Circulating DNA Kit (protocol: circDNA3 CR22161 ID2686). We used 4ul of the resulting DNA for ddPCR assays.

We designed two probes targeted at the wildtype (reference) allele and the R206C allele (alternative). Our ddPCR experiments therefore resulted in an observation of the number of reference alleles and a number of alternative alleles, corresponding to the number of positive droplets on either channel-1 or channel-2, which was measured by the level of fluorescence.

To determine the most likely maternal and fetal genotype (G) given an observation (O) from a ddPCR experiment, we proceed as follows.

We first define *O* as a 2-tuple describing the ddPCR observation of allele counts, e.g. (5,11) refers to an observation in which the reference allele was observed 5 times, while the alternative allele was observed 11 times. *O*_0_ refers to the reference allele, and *O*_1_ refers to the alternative allele.

The fetal and maternal genotype G are represented by a 3-tuple describing the maternal and fetal genotype, in which G_0_ refers to the inherited maternal allele, G_1_ refers to the non-inherited maternal allele and G_2_ refers to the inherited paternal allele. We use 0 to denote the reference allele and 1 to denote the alternative allele, (0,1,1) would present the case where both mother and fetus are heterozygous, but the alternative allele was transmitted through the paternal genome.

### Now the likelihood of O given G can be derived as follows

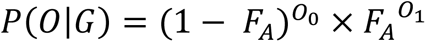

Where, *F*_A_ is the expected frequency of the alternative allele under *G*. Which can be derived from:

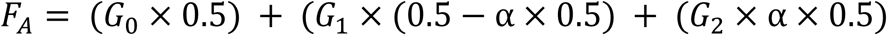

Where α represents the fetal fraction, which can either be inferred from the Y-chromosome in case of male-bearing pregnancies or a fragmentomics predictor. Alternatively, we chose to use a default value of 0.05.

To simplify, we assumed a uniform prior probability (P(*G*)) across all 8 possible genotypes (0.5^3^). The estimated minor allele frequency of a variant in the population could be used here as well for a more accurately model, but this did not change our results.

The marginal probability (P(O)) can be obtained by summing over the product of prior and likelihood for all possible genotypes.

Now, the posterior probability (P(G|O)) of the maternal and fetal genotype at a bi-allelic variant can be derived from Bayes formula:

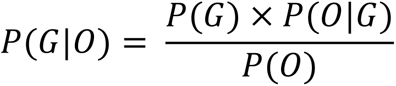

Iterating over all 8 possible genotypes can determine the most likely genotype. The most likely maternal genotype (the number of alternative alleles in the maternal genome) then corresponds to G_0_+G_1_, while the most likely fetal genotype corresponds to G_0_+G_2_.

Note that this model only holds for non-surrogate pregnancies, and in cases where both mother and fetus are heterozygous, this model cannot distinguish whether the allele was inherited through the maternal or paternal genome as *F*_A_ would be identical.

#### SUPPLEMENTAL INFORMATION TITLES AND LEGENDS

Table S1 Genome-wide significant loci from cfDNA GWAS, related to Figures 1 and 2 and Table 1.

Table S2 Effect of R206C on cleave-site motifs and validation cohort, related to Figure 3.

Table S3 Results of LD-score regression: genetic correlations and partitioned heritability, related to Table 1.

